# Early changes in the gut microbiome among HIV-infected Individuals in Uganda initiating daily TMP/SMX

**DOI:** 10.1101/2024.10.07.24315002

**Authors:** Carolyne Atugonza, Adrian Muwonge, Christine F. Najjuka, David P. Kateete, Eric Katagirya, Savannah Mwesigwa, Benon Asiimwe

## Abstract

Daily cotrimoxazole (TMP/SXT) prophylaxis is part of the HIV treatment package for all new HIV-infected individuals in Uganda. Although this treatment has shown reduced morbidity and mortality in HIV, it remains controversial due to its contribution to developing antibiotic-resistant bacteria. Moreover, the effects of daily use of a broad-spectrum antibiotic on the gut microbiome remain unknown.

To study the early effects, we analysed shotgun metagenome sequence data from stool samples of five newly HIV-infected individuals initiating TMP/SXT prophylaxis longitudinally for the first 30 days of treatment. Using shotgun metagenomics sequencing, we generated both taxonomic and functional profiles from each patient and compared gut microbial changes Pre-TMP/SXT and post-TMP/SXT on Day 5, Day 14, and Day 30.

Daily TMP/SXT prophylaxis resulted in a shift characterised by an enrichment of *Prevetollea* and *Ruminococcus* genera members and the depletion of *Lactococcus* and *Bacteroides* genera members. Furthermore, these microbial shifts were associated with changes in the functional profile revealed by a differential abundance of pathways of amino acid metabolism, carbohydrate metabolism, and nucleotide biosynthesis linked to members of the Bacteroidaceae and Enterobacteriaceae families.

TMP/SXT daily prophylaxis in HIV-infected individuals is associated with dramatic changes in microbial composition and functional profiles; however, other factors such as Age, Gender, HIV clinical stage, and ART regiment are at play. Further investigation is needed to examine the implication of these shifts on clinical management and outcomes among HIV patients.

## Background

The human gastrointestinal tract (GIT) is a complex system made up of a large number of microorganisms. The GIT comprises bacteria, archaea, viruses, parasites, and fungal species(Lu *et al*., 2018), collectively known as ‘gut microbiota. The gut microbiota plays essential physiological functions, including developing and maintaining host immunity, aiding digestion, and helping control pathogenic bacteria (Kau *et al*., 2011; Thaiss *et al*., 2016; Liu *et al*., 2017).

However, during the Human immunodeficiency virus (HIV) infection, this microbial community is highly destabilised, as seen by a reduction in microbial diversity and loss of beneficial bacteria in a phenomenon known as dysbiosis, often characterised by reduced abundances of Bacteroides and increased abundance of genus Prevotella (Zevin *et al*., 2016). Such altered gut integrity due to HIV infection modifies the gut mucosal barrier resulting in a change in the gut microbiota and a shift to proinflammatory or potentially pathogenic bacterial populations causing opportunistic infections (Dillon, Frank, and Wilson, 2016). Such infections continue to cause significant mortality and morbidity worldwide in HIV-infected individuals. In high-income countries, such conditions are limited, with Pneumocystis jiroveci pneumonia being the most prevalent (Kaplan *et al*., 1996). On the other hand, low-income countries have a high occurrence of opportunistic infections, including parasitic enteritis and bacterial diseases like tuberculosis and salmonella, mainly affecting HIV-infected individuals (Holmes *et al*., 2003; Lowe *et al*., 2013).

Cotrimoxazole treatment or prophylaxis against several of these opportunistic infections is offered alongside antiretroviral therapy (ART). Cotrimoxazole, a fixed-dose combination of sulfamethoxazole and trimethoprim (TMP/SMX), is abroad spectrum antimicrobial agent that targets a range of aerobic gram-positive and negative organisms, fungi, and protozoa (D’Souza *et al*., 2020). With recommendations from the WHO, TMP/SXT prophylaxis is incorporated into the health care package for all HIV-exposed individuals(WHO, 2014), particularly for areas where malaria and bacterial infections are highly prevalent. In compliance with WHO recommendations, Uganda offers TMP/SXT preventative therapy for a particular category of patients, including all people living with HIV initiating ART, Pregnant and breastfeeding women, children and adolescents aged 15 and below, and all patients suspected to have treatment failure(of Health and Republic of Uganda, 2016). Although WHO guidelines denote changes in the gut microbiome caused by TMP/SMX, studies and details of these changes remain limited, especially in Uganda (WHO, 2014).

Notably, a knowledge gap remains in the understanding of the early effects of TMP/SMX on the gut microbiome in HIV patients. Using the nationally scheduled clinical visit dates in the HIV care program in Uganda; we examined the 30-day longitudinal impacts of TMP/SMX initiation during HIV treatment on the Gut microbiome by utilising shotgun metagenomics. Metagenomics sequencing allows comprehensive sampling of all genes in organisms in a sample (Mardis, 2008). It enables the evaluation of bacterial diversity and abundance of microorganisms, which are missed by cultivation-based methods. Moreover, functional metagenomics can identify novel functional genes, microbial pathways, and dysbiosis of the intestinal microbiome and determine interactions and co-evolution between microbiota and host. This study reports on the gut microbiota taxonomic and functional profiles examined in the study cohort.

## MATERIALS AND METHODS

### Study Population and Sample

This prospective, preliminary study was conducted to study the effects of TMP/SMX on the gut microbiome in 5 adult HIV-infected patients. The School of Biomedical Sciences Research and Ethics Committee of Makerere University (IRB No.0006291) approved the study. We enrolled newly diagnosed HIV-infected individuals reporting at the Makerere Joint AIDS Project (MJAP) ISS clinic at Mulago Hospital from February 2019 to May 2019. Critical inclusion criteria are as follows; (i) Had not received any antibiotic treatment in the last 30 days (ii) No comorbidities with infectious agents. All participants provided written informed consent before study enrollment. Upon enrollment, all participants continued with the recommended HIV care package freely provided by the HIV Clinic, including the prescribed ART regimen and TMP/SMX. For TMP/SMX, the prescription was provided based on national guidelines structured according to a patient’s weight; <5kg 120mg, 5-14.9kg 240mg, 15-29.9kg 480mg, ≥30kg 960mg. Fresh stool samples were collected from participants during their clinic visit at four different time points, including a baseline sample before TMP/SMX treatment, Day 5, Day 14, and Day 30 after TMP/SMX to study the early effects of TMP/SMX treatment. Samples were frozen within 4-6 hours of collection and stored at -80°C until analysis. Twenty stool specimens were collected from the five patients during the study.

### Stool DNA Extraction and sequencing

According to the manufacturer’s instructions, total bacterial DNA was extracted using the Powerlyzer DNA extraction kit (Qiagen Inc., Germany). Total DNA concentration and purity were assessed by absorbance on a Nanodrop N1000 (Thermo Fisher Scientific, Carlsbad, CA, USA) by measuring the A260/A280 ratio before storage at -20°C. DNA libraries were prepared using Illumina Truseq Nano DNA Library Preparation kits, quantified, and normalised before 20 samples were pooled per sequencing lane. Shotgun metagenomic sequencing was performed using the DNBseq BGI platform at the BGI institute, China, with a sequencing depth of 5 Gbps and an insert size range of 275-450 bp per sample.

### Sequence quality assessment

The raw sequencing data were processed using METAWRAP(Uritskiy, DiRuggiero and Taylor, 2018) pipeline to remove low-quality reads, adapters, and human DNA contamination. In brief, reads were trimmed using a length cut-off of 30; subsequent, trimmed reads were then screened against adapter sequences and the human genome sequence with a 90% identity cut-off. After filtering, a mean of 21 million sequences was obtained for every metagenome sample.

### Taxonomic and Functional Profiling

For each sample, taxonomic profiling for cleaned reads was obtained using MetaPhlAn2 Software (Segata *et al*., 2012) and functional profiling using HUMAnN (Franzosa *et al*., 2018). Sequence data were deposited in the NCBI Sequence Read Archive database.

### Statistical Analysis

All statistical analyses were performed using the R program version 3.5.1. Alpha diversity was calculated using normalised counts, and the Kruskal-Wallis test determined the significance. Beta diversity was determined using the Bray-Curtis distance matrix. Principal component analysis (PCoA) was performed on the calculated distance matrixes, and PERMANOVA or Adonis calculated significance with 999 permutations. PCoA plots and confidence ellipses were generated by ggplot and ggpubr version package in R. All P values <.05 were considered statistically significant for all tests.

## RESULTS

### Clinical and demographic characteristics of the study cohort

Five HIV-infected Ugandan subjects were enrolled in the study at the MJAP HIV clinic at Mulago Hospital. Participants were primarily female, with a median age of 31 and a median body mass index (BMI) of 23.6 kg/m2. During admission to the clinic, patients were also classified according to clinical stages of the WHO (Weinberg and Kovarik, 2010): Stage 1: 3 patients; Clinical Stage 2: 1 patient; Clinical Stage 3; 1 patient, and no patients with clinical stage 4 were enrolled. CD4 count data was also obtained during the enrolment of the study. Four participants had a CD+4 T-cell count of ≥ 500 cells/μL, with one participant having a 200-500 cells/μL count and 4 participants ≤ 200 cells/μL at the time of enrolment. All the participants were started on antiretroviral treatment and cotrimoxazole prophylaxis treatment. Three participants were started on TDF/3TC/DTG regimen, while two were started on TDF/3TC/EFV regimen (Table 1).

**Table 1.**
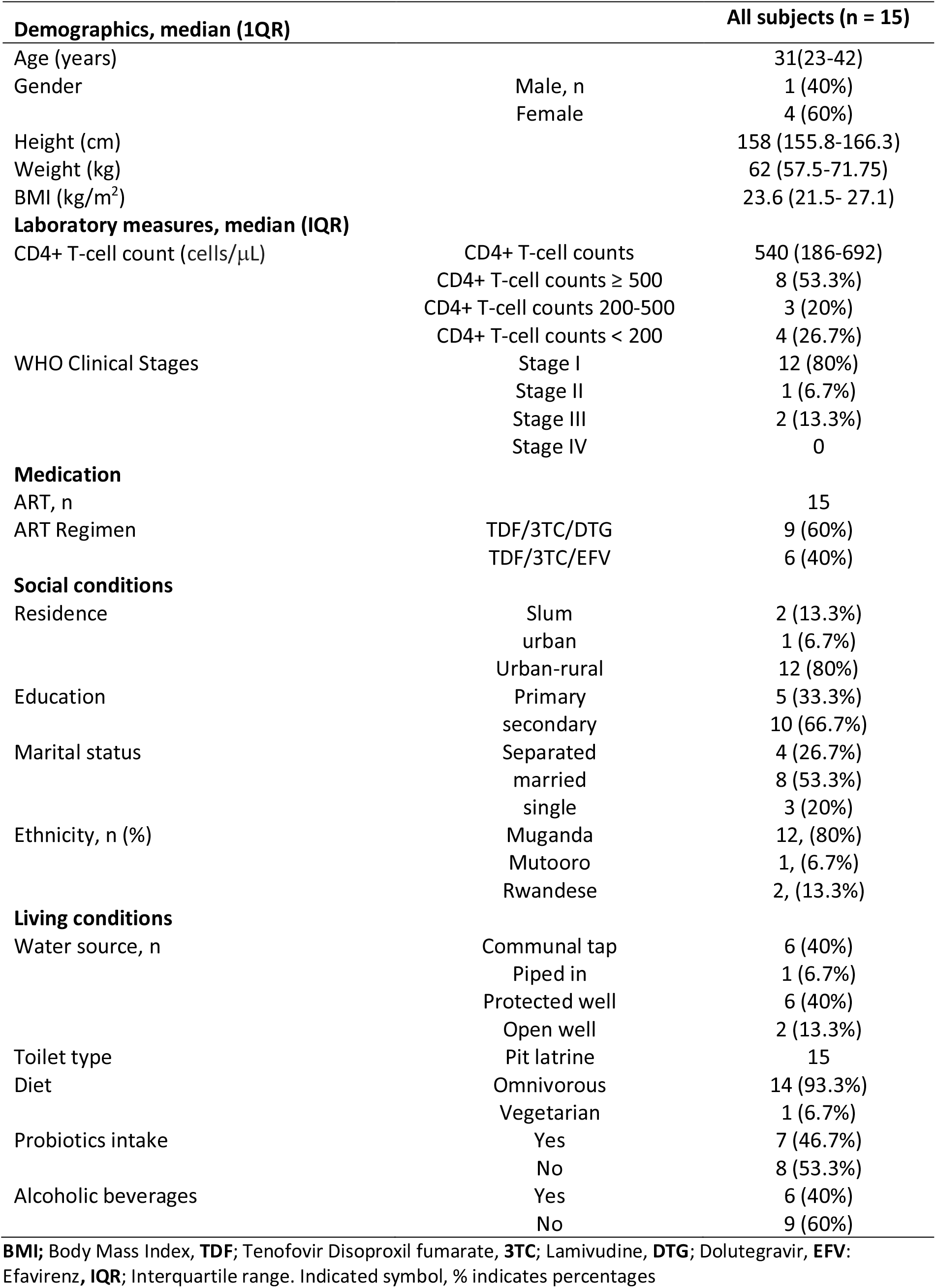
Baseline characteristics of HIV patients enrolled in the study cohort.

### Changes in microbial diversity along the treatment timeline among HIV patients

To investigate the impacts of Early ART treatment with TMP/SMX on the gut microbiome, we longitudinally compared sampled gut microbiome profiles of HIV-infected individuals. Twenty stool samples from 5 HIV-infected individuals were collected longitudinally for up to 4 consecutive weeks.

To examine whether there was an alteration in bacterial composition, we compared alpha diversity before (Pre-TMP/SMX) and after TMP/SMX initiation (Post-TMP/SMX). Post TMP/SMX was associated with decreased α-diversity (*P* = 0.053, Shannon; P = 0.053) (Figure 1A). When we compared alpha diversity at the different time points, a difference was observed between Baseline and Day 30 after TMP/SMX initiation (Kruskal-Wallis, P =0.056) (Figure 1B). As expected, alpha diversity was lower in samples on Day 5, Day 14, and Day 30 after TMP/SMX administration. Only Age and Gender were associated with significant alpha diversity compared to other metadata. (Supplementary Figure 1S).

**Figure 1.**
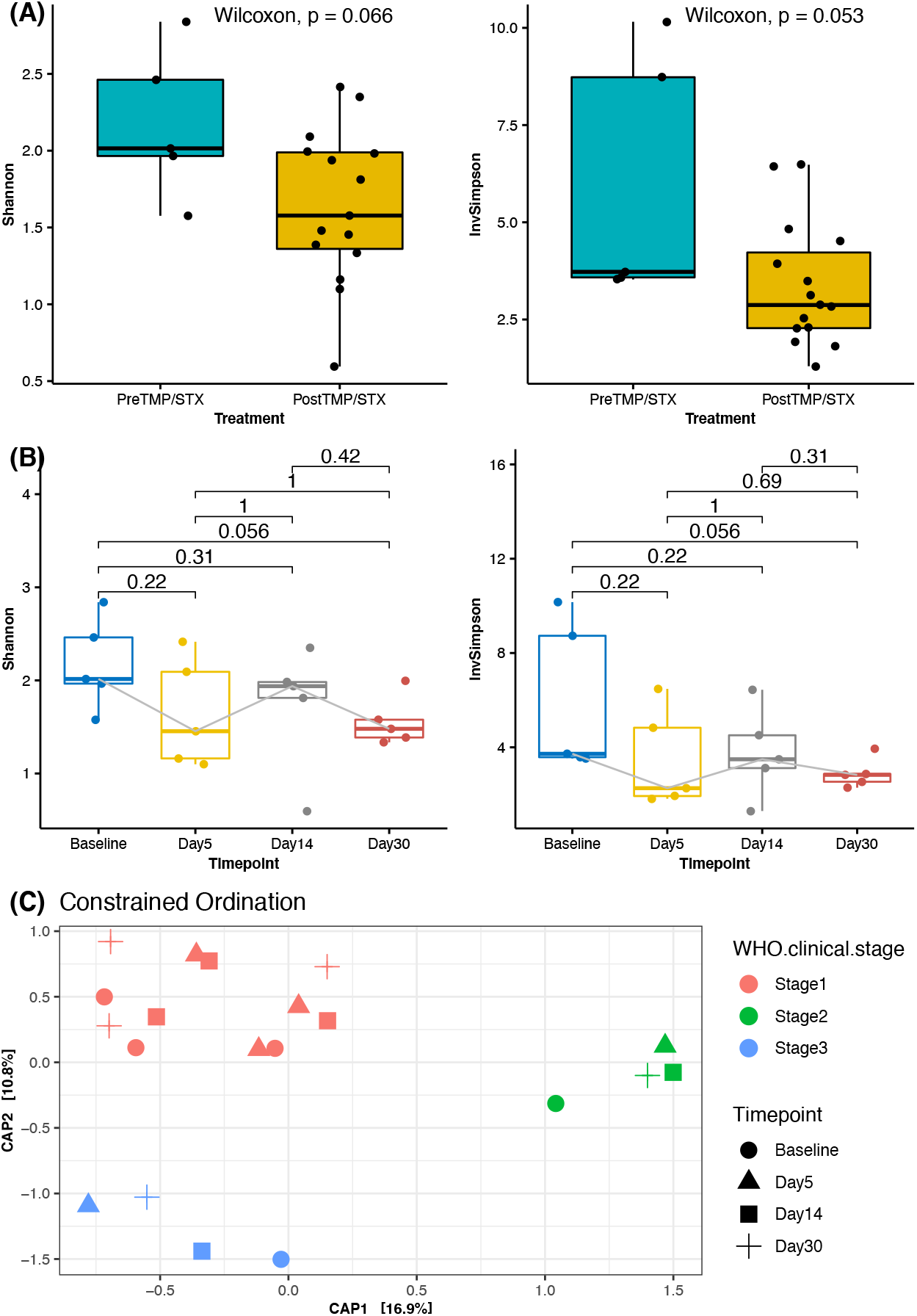
Signs of Dysbiosis in HIV-infected individuals initiating TMP/SXT. (A) Alpha diversity (Shannon Index and InvSimpson) before and after TMP/SMX initiation. (B) Comparison of alpha diversities between the different time points showing a difference between Baseline and Day 30. (C) Principal coordinates Analysis (PCoA) of Bray-Curtis distances at the different time points showing clustering patterns according to HIV clinical stages.

The differences between pre and post-TMP/STX suggest an association in gut microbial changes; however, the PCoA using Bray-Curtis distance shows no significant structural changes since no significant clustering at phyla and genera at the different time points (Linear mixed model, Bray-Curtis, R2=0.084, P = 0.941) were observed. On the other hand, the HIV stages were associated with gut microbial structure, explaining up to 36.7% of the structural variation (Adonis; *P* < 0.05, R^2^ = 0.41186). Similarly, treatment regimen types were associated with gut microbiome differences, explaining 26.9% of structural variation (Adonis; *P* < 0.05, R^2^ = 0.30186) (Figure 1C). Further, multivariable PERMANOVA analysis showed age explained 36%, gender 14 variation in the microbiota composition, indicating that they may be drivers of distinct microbial community composition. In addition, individuals tended to cluster together, suggesting the existence of distinct microbiota unique to individuals despite the same treatment. Other tested variables did not significantly contribute to the variability in microbiota composition (Table 2)

**Table 2.** Permutational multivariate analysis of variance (PERMANOVA) table for microbial community composition from study groups before and after TMP/STX and associated metadata.

| Variable | F.Model | R<sup>2</sup> | Pr(>F) |
| --- | --- | --- | --- |
| Treatment groups (Pre TMP/STX and Post TMP/STX) | 1.225 | 0.064 | 0.282 |
| Timepoints | 0.496 | 0.085 | 0.918 |
| CD4 count | 1.180 | 0.061 | 0.296 |
| BMI | 1.022 | 0.054 | 0.378 |
| Age | 10.246 | 0.363 | 0.001* |
| Gender | 3.000 | 0.143 | 0.021* |
| HIV clinical Stage | 4.922 | 0.367 | 0.001** |
| ART Regimen | 6.636 | 0.269 | 0.002** |

PERMANOVA assessed the significance of associations between Bray-Curtis dissimilarity and the different variables. P-values are based on 999 permutations. \* represents significant P values. ( $P < 0.05$ )

### TMP/SMX treatment and associated changes in gut taxonomic profiles

Here we sought to examine how the microbiota community varied at different time points from the Baseline to Day 5, Day 14, and Day 30. Bacteroides were the most abundant gut phyla among the samples, followed by Firmicutes, Proteobacteria, and Actinobacteria (Figure 2A). A comparison of bacteria at the family level demonstrated abundance in Bacteroidaceae, Prevotellacea, Ruminococcaceae, and Lachnospiraceae (Figure 2C). The top genera included *Bacteroides, Prevotella, Eubacterium, Parabacteroides, Feacalibacterium, Ruminococcus, and Roseburia*, with *Prevotella* and *Bacteroides* having the highest abundance (Figure 2B).

**Figure 2.**
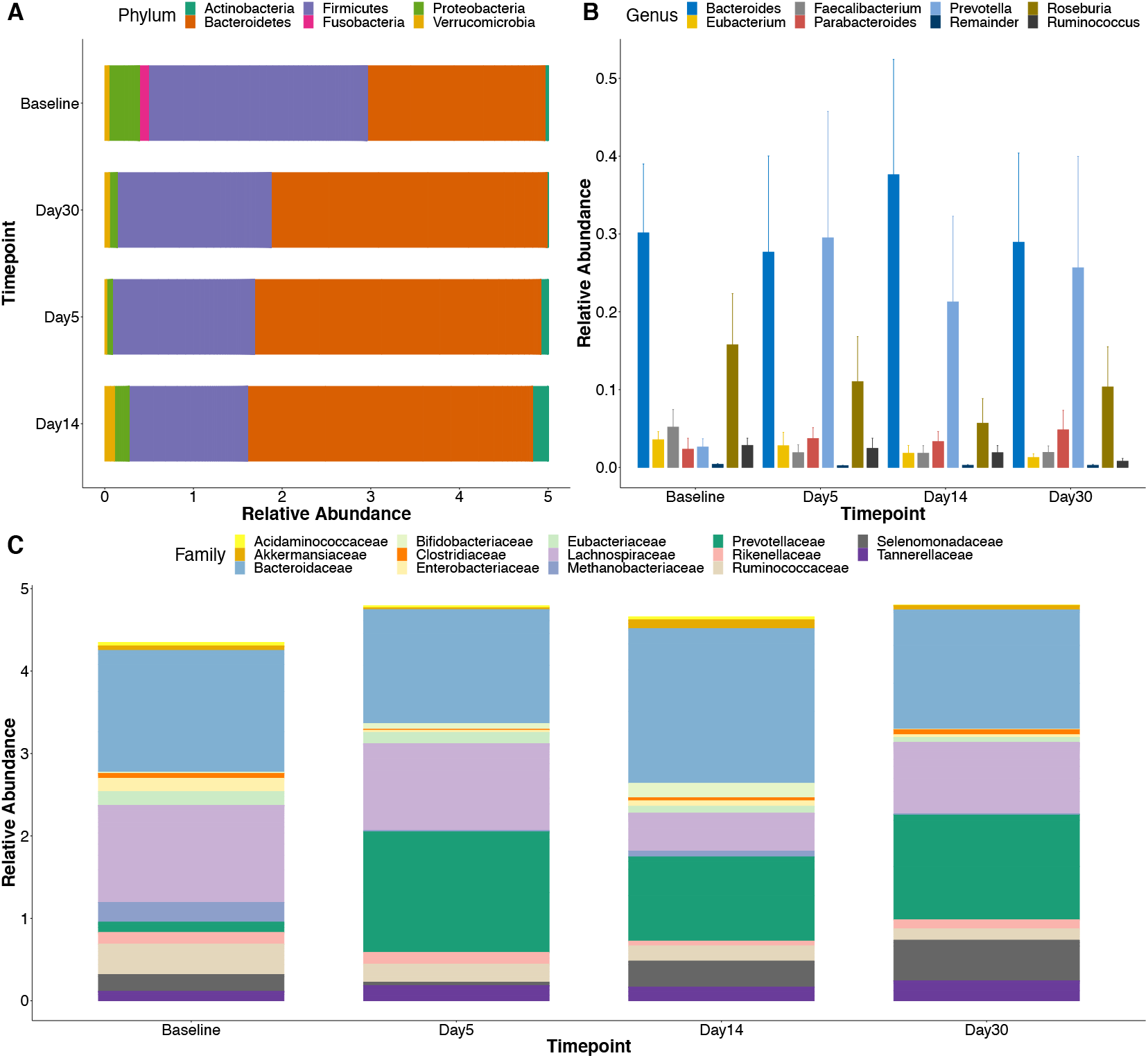
Early Taxonomic changes in the gut microbiota of HIV-infected individuals initiating TMP/SXT treatment are dominated by Bacteroidetes. Bacteria taxa were identified in stool samples of 5 HIV-infected individuals initiating TMP/SXT treatment and followed up for 30 days using Shotgun metagenomics sequencing. (A) A stacked bar plot analysis showing the top 6 phylum abundances at each time point. (B) Relative abundance of the top genus at the different time points. (C) Comparison of the Abundances of top phyla showing Protobacteria having a statistical difference at the different time points. # Statistical difference was calculated using the Kruskal-Wallis test.

We further examined the differential abundance of genera with the progression of time using DESeq2. By accounting for patient differences, results revealed shifts in taxa abundance at the different visit dates. From Baseline until day 30, we observed an enrichment of the Prevotella genus. On day 5, a depletion of the Lactococcus, Clostridium, Ruminococcus, Fusobacterium, and Klebsiella genera was observed. On day 14 and Day 30, a similar trend as on Day 5 was observed with an additional depletion of the Bacteroides genus (Supplementary Table S1).

Further analysis was conducted to examine any changes at the species level. Special abundances at the different time points showed many shifts in OTU abundance that were statistically (FDR < 0.05) significant. Using the Baseline as a reference, two species, *Ruminococcus gnavus* and *Prevotella copri*, were significantly enriched on day 5, whereas day 14 revealed Prevotella copri and Bacteroide pectinophilus enrichment. On day 30, a similar trend as on day five was observed (Figure 3). Other species of *Bacteroide caccae, Bacteroides pectinophilus, Barnesiella intestinihominis*, Clostridium_sCA413, *Fusobacterium Mortiferum, Holdemanella biformis, Klebsiella variicola, Lactococcus tauri, RuminococcusCA488* had a significant decrease throughout treatment and did not return to baseline levels. The results reveal a major compositional shift in the microbiota on Day 14 and Day 30 with a significantly increased fold change (FC>10, FDR < 0.05) of some species suggesting a possible impact by antimicrobial treatment.

**Figure 3.**
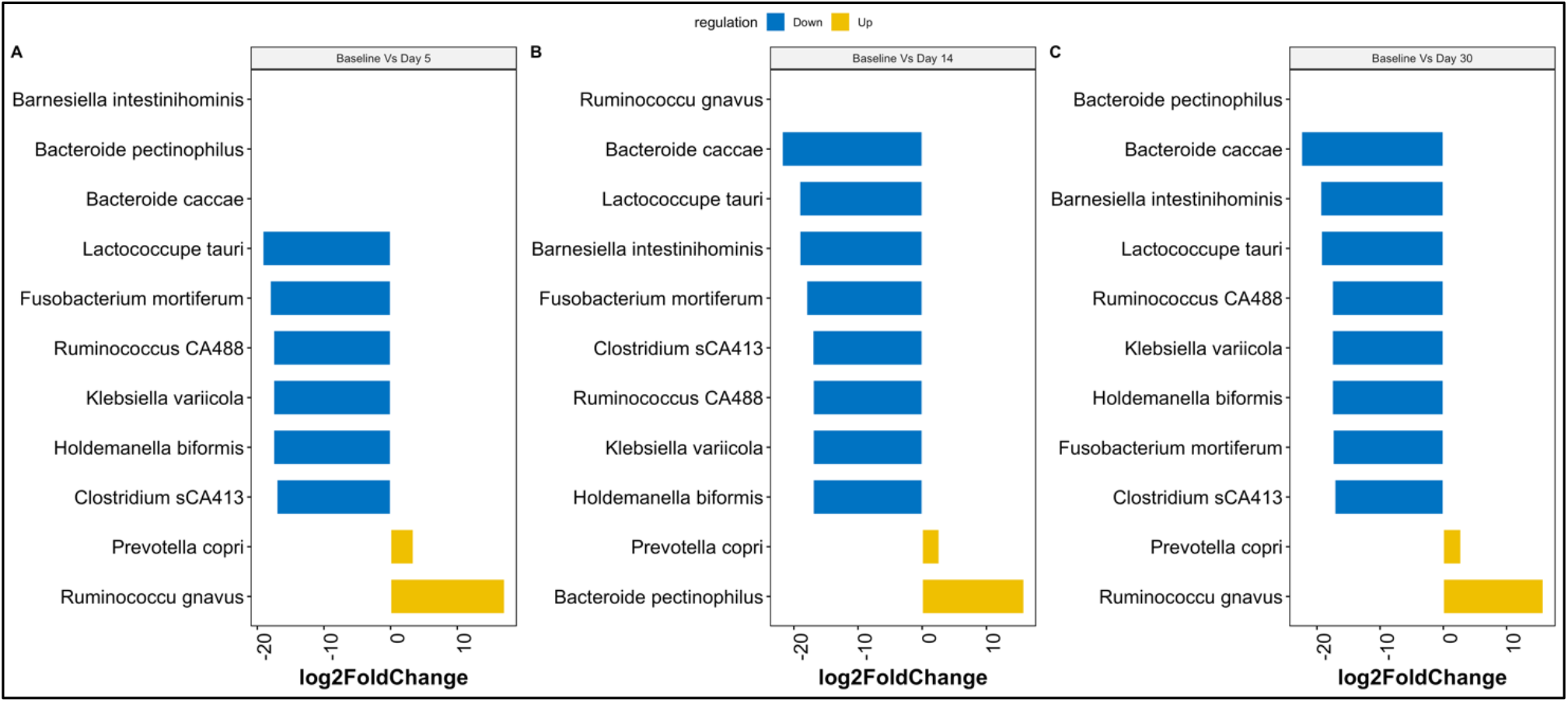
Changes in taxonomic abundance in the gut microbiome. Fold change scores determined by DESeq2 analysis reveal differences in the abundance of key Species between different time points. Significant group differences were corrected with Benjamini–Hochberg False Discovery Rate p FDR < 0.05.

### Changes in microbial functional profiles following TMP/STX

Next, we further investigated the functional profiles in the gut microbiome during TMP/STX uses to obtain pathway abundance HUMANn2 pipeline. This functional profiling analysis revealed 400 pathways and heatmap analysis of the top 30 pathways across all the samples (Figure 4). Diversity analysis revealed no significant differences between the pathways based on the different time points with Shann (Supplementary Figure 2S).

**Figure 4.**
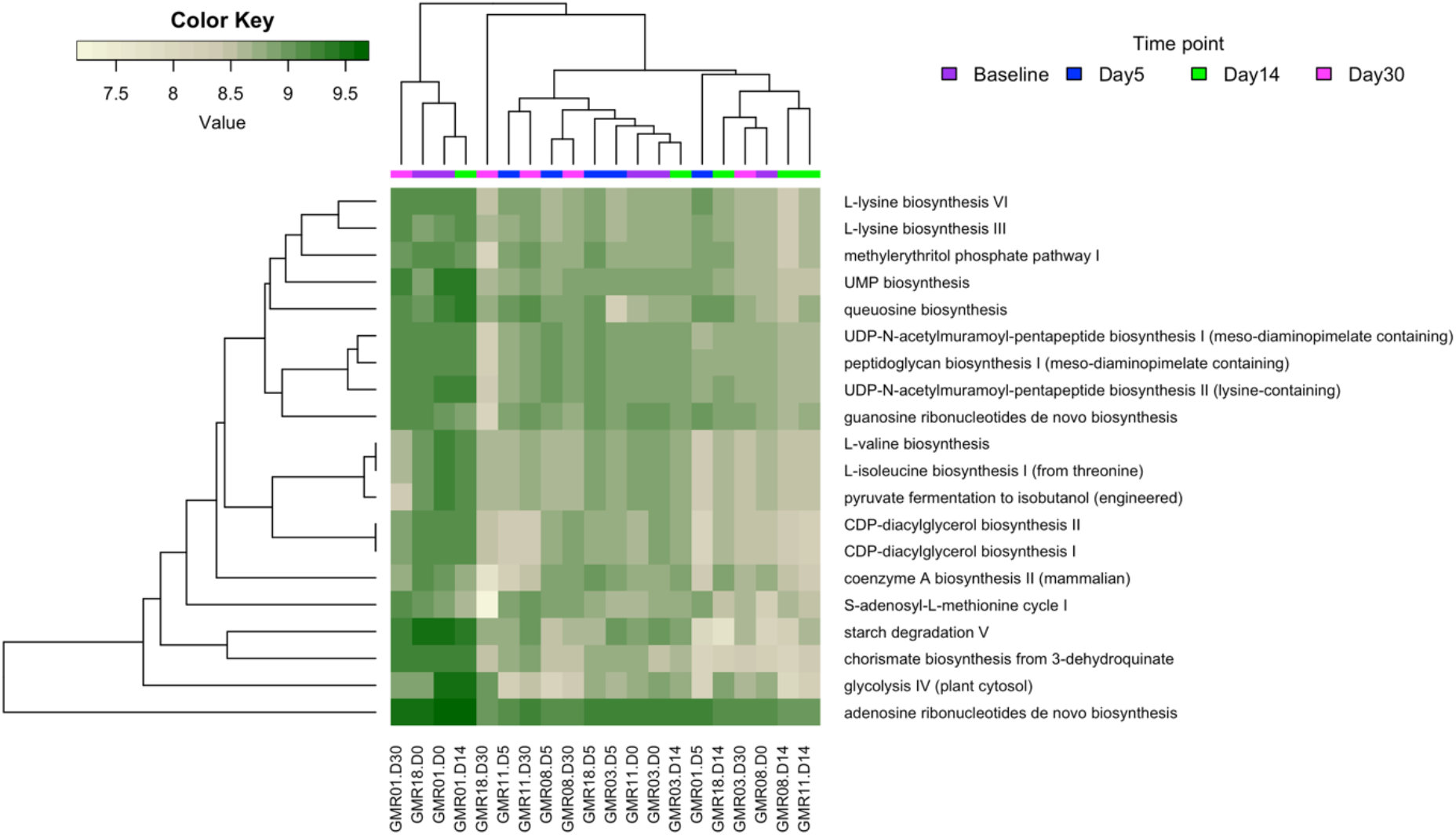
Inferred functional profile of the Gut microbiome during TMP/SXT administration for 30 days. Heatmap of the top 30 most abundant pathways in the individual samples as recognised by HUMAnN2. Samples were hierarchically clustered using Bray-Curtis dissimilarity for the pathways and colour-coded based on the time point shown in the image.

**Figure 5.**
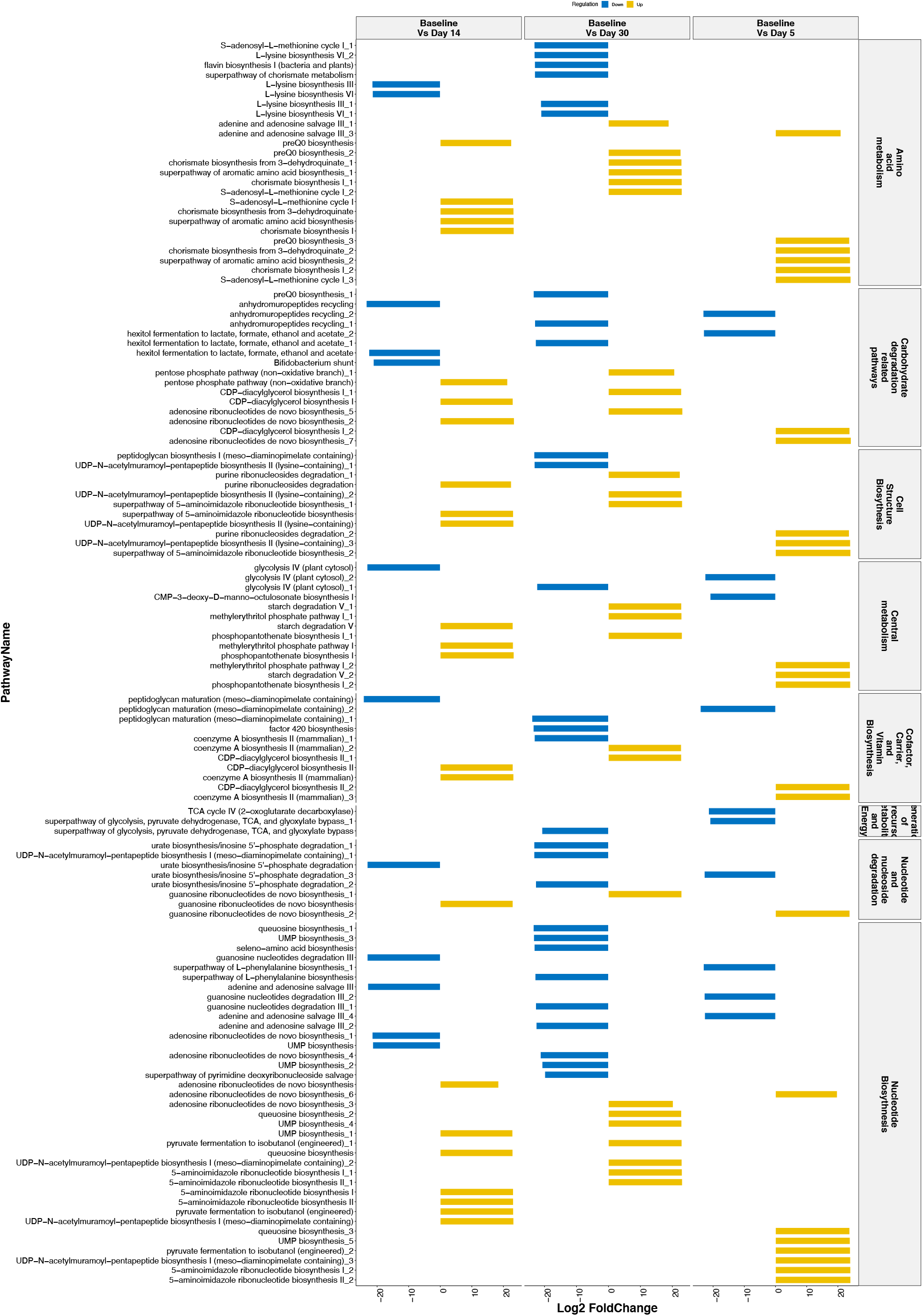
Differential Abundance analysis reveals fundamental pathway changes during TMP/SXT initiation in HIV-infected individuals. Bar plot of Identified pathways from HUMANn2 showing DESeq2 log2 fold change values for comparisons at the different time points using Baseline as a reference. Pathways represented were grouped into different metabolic classes. Duplicated pathways were reported as two or more bacteria attributed to the pathway’s contribution.

In further analysis, we investigated how the metabolic activity changed during treatment. Here, we studied how the pathways changed at each time point about the pathways observed at the baseline time. Using DESeq2 (Figure 4), changes on Day 5 revealed the enrichment of Purine Nucleotide synthesis pathways (Adenosine ribonucleotide de novo biosynthesis and 5-aminoimidazole ribonucleotide biosynthesis pathways) and depletion of Cell wall biosynthesis pathways (peptidoglycan maturation) and Purine Nucleotide Degradation pathways (adenosine nucleotides degradation II and guanosine nucleotides degradation III). On Day 7, enriched pathways included Purine Nucleotide pathways (Adenosine ribonucleotide de novo biosynthesis), Aromatic compound biosynthesis (Chorismate biosynthesis I and super pathway of aromatic amino acid biosynthesis), and Carrier Biosynthesis (Phosphopanthothenate biosynthesis I). Cell wall biosynthesis pathways (peptidoglycan maturation), Sugar derivative degradation (Anhydromuropeptides recycling), and central metabolism pathways (Glycolysis IV) were the most depleted.

Similarly, by Day 30, Purine Nucleotide synthesis pathways (Adenosine ribonucleotide de novo biosynthesis and 5-aminoimidazole ribonucleotide biosynthesis) were enriched. Top depleted pathways included Carrier Biosynthesis pathways (flavin biosynthesis II and factor 420 biosynthesis), Cell wall biosynthesis pathways (peptidoglycan maturation), and aromatic compound biosynthesis (Chorismate biosynthesis I). A summary of depleted and enriched pathways is represented in a table (Supplementary Table S2).

### Inferring functional taxonomic communities

Data were further analysed to understand which species contributed to the enrichment or depletion of significant functional pathways. The important discriminative pathways enriched between Baseline and day 5 (PWY) were mainly contributed *by Butyrivibrio crossotus*, one pathway (adenine and adenosine salvage III) by *Lachnospiraceae bacterium*, and one pathway (adenosine ribonucleotides de novo biosynthesis) by *Acidaminococcus fermentans*. One species, Shigella sonnei, contributed *to all depleted pathways*. (Supplementary table). Similarly, on Day 14, enriched pathways were contributed mainly by *Butyrivibrio crossotus*, and depleted pathways were contributed by *Shigella_sonnei* and *Bacteroides stercoris*. By Day 30, enriched pathways were primarily contributed by *Butyrivibrio crossotus*, and depleted pathways were contributed by three species *Shigella sonnei, Bacteroides stercoris, and Barnesiella intestinihominis*. These results are consistent with our previous results that showed a high representation of the family *Lachnospiraceae, Bacteroidacea, and Barnesiellacea* in the gut microbial abundance (Supplementary Figure 3S).

## Discussion

In this study, we investigated the early effects of daily TMP/SXT on the gut microbiome of HIV-infected individuals. Patients were followed up for 30 days with samples collected at the Baseline before treatment initiation (Baseline) and at day 5, day 14, and day 30 to study the early effects of TMP/SXT on the gut microbiome taxonomic and metabolic profiles.

### Shifts in functional profiles associated with TMP/SXT

Results revealed a varied functional profile at the different time points during treatment with pathways involving Amino acid metabolism, Carbohydrate metabolism, Nucleotide biosynthesis and degradation, cell structure, and cofactor/ carrier/ Vitamin biosynthesis during TMP/SXT treatment.

From Baseline to day 30, we found an enrichment of pathways involved in amino acid metabolism in the gut microbiota. In particular, pathways of chorismate biosynthesis, aromatic amino acid biosynthesis, L-methionine, and adenine and adenosine salvage were upregulated throughout treatment. In contrast, continuous depletion of L-lysine and flavin biosynthesis was observed during treatment. Pathways involved in amino acid metabolism play a role in shaping the response of immune cells (O’Neill, Kishton and Rathmell, 2016). However, Lysine depletion would be of concern as a significant percentage of circulating lysine in the host has been reported to be derived from microbial sources (Metges, 2000).

In addition, we observed the enrichment of the purine nucleotide synthesis pathways, Adenosine ribonucleotide de novo biosynthesis, and 5-aminoimidazole ribonucleotide biosynthesis pathways throughout treatment (Day 30). These pathways involve nucleic acid synthesis, cell signalling, and energetic homeostasis. 5-aminoiamidazole ribonucleotide is the crucial intermediate for purine nucleotide biosynthesis of the gut microbiota (Sheng *et al*., 2021). The high abundance of these pathways may indicate significant bacterial metabolic activity during growth. In addition, we identified differential abundances of pathways involved in central metabolism and pathways in cofactor, carrier, and vitamin biosynthesis. Pathways altered were responsible for the production of energy and nucleotides. Moreover, this is consistent with studies that have shown bacteria respond to the stress of antibiotics through the diversion of ATP from core metabolic processes to drive the maintenance of ion gradients (Stokes *et al*., 2019).

Important to note that members of Firmicutes majorly contributed functional roles among bacterial taxa for enriched pathways; *Butyrivibrio crossotus, Lachnospiraceae bacterium_2_1_58FAA, Eubacterium siraeum*, and *Acidaminococcus fermentans*. Members of Clostridia are known for their ability to participate in vital metabolic processes, including amino acid fermentation and polysaccharide degradation (Dai, Wu and Zhu, 2011). These taxa are also members of butyrate producers. The association of these observed bacteria suggests that antibiotic exposure promotes an abundance of butyrate-producing *Clostridium* members. On the other hand, depleted pathways were contributed by Proteobacteria member *Shigella sonnei* and Bacteroidetes members; *Bacteroides stercoris* and *Barnesiella intestinihominis*. Further study is needed to examine this microbial functional signature and its role during TMP/SXT.

Subsequently, these findings raise an important consideration of how taxonomic shifts caused by TMP/SXT pressure may simultaneously offer valuable insight into changing active communities. Alterations of such pathways may have important physiological significance due to TMP/SXT impact on the gut, given that the products of such metabolisms, such as short-chain/branched-chain fatty acids and biogenic amines, can cause changes in the intestinal environment (Tremaroli & Bäckhed, 2012); however, more research is warranted. Although these results contribute to understanding the early impacts of TMP/SXT in the gut microbiota, it would be essential to identify which metabolic profiles are genuinely relevant and are a consequence of antibiotic treatment. Moreover, this would provide an opportunity to target which pathways are associated with TMP/SXT treatment to reduce the impact of gut microbiota dysbiosis.

### Structural and compositional shifts associated with TMP/SXT

Next, we analysed how TMP/SXT impacted the gut microbial composition. We observed compositional shifts were mainly characterised by an abundance of Bacteroidota, Firmicutes, Proteobacteria, and Actinobacteria at the Phylum level in the gut microbiome of Patients initiating TMP/SXT therapy similar to other studies (Dillon *et al*., 2014; Monaco *et al*., 2016; Vujkovic-Cvijin and Somsouk, 2019; Parbie *et al*., 2021). We did not observe significant phylum level changes with time in the study. At the family level, we reported an abundance in Bacteroidaceae, Prevotellaceae, and Lachnospiraceae, whereas *Bacteroides, Prevotella*, and *Roseburia* dominated the genus level, similar to a study by Machiavelli et al. (Machiavelli *et al*., 2019). The composition shifts at the species level show enrichment of species belonging to *Prevetollea* and *Ruminococcus* genera and depletion of species belonging to Lactococcus and Bacteroides genera. Here we note an enrichment in *Ruminnococcus gnavus and Prevotella copri. Ruminnococcus gnavus* is an anaerobic, gram-positive member of the gut microbiome. *Ruminococcus* are associated with protective and disruptive roles in the gut by immune modulation through the production of Fatty acid chains (Flint *et al*., 2008) or its proinflammatory actions as reported in IBD (Png *et al*., 2010). Moreover, the enrichment with the genus Prevotella has been exhibited in other HIV studies (Vujkovic-Cvijin *et al*., 2013; Lozupone *et al*., 2014; Mutlu *et al*., 2014). Similarly, a study by Kaur et al. 2018 also observed elevated levels of *Prevotella copri* and its potential inflammatory role in the HIV disease (Kaur *et al*., 2018). Additionally, we reported a depletion in *Bacteroides caccae* and *Lactococcus taur*i. A said low abundance in several species of the genus Bacteroides has an anti-inflammatory function by promoting T regulatory cell reactivity, and these microorganisms have also been associated with increased inflammation (Pérez-Santiago *et al*., 2013; Jiang *et al*., 2017; Lee *et al*., 2018; Neff *et al*., 2018; Xie *et al*., 2021). Decreases in commensals, including *Lactococcus*, have been reported during HIV infection (McHardy *et al*., 2013; Mutlu *et al*., 2014). *Lactococcus* is also associated with potential benefits to the host by producing bacteriocins and organic acids (Yan Yang *et al*., 2016). Overall, trends in microbiota have been reported with a signature increase in Prevotella and a decrease in Bacteroides in HIV-infected individuals.

We also found a microbial compositional shift after TMP/SXT initiation associated with a significantly decreased diversity compared to pre-antibiotic use and a microbial shift associated with Age, HIV clinical stage, and ART regimen use. Previous studies support our observation that TMP/SXT use causes compositional changes in the gut during its use(ref). In contrast, a study by Monaco et al. (Monaco *et al*., 2016) on HIV individuals using TMP/SXT did not reveal significant changes in the alpha diversity of studied individuals. Important to note is the difference in study design as this study was conducted on individuals on long-term use of TMP/SXT. Thus, although early effects of TMP/SXT may initially impact the alpha diversity, these effects may be resolved over time, as shown in other studies (Raymond *et al*., 2016; Palleja *et al*., 2018; Shaw *et al*., 2019). Similarly, D’zoula et al. (D’Souza *et al*., 2020) found that infants on TMP/SXT had a reduced β-diversity compared to their non-TMP/SXT counterparts.

Although the CD4+ T cell population is highly depleted during HIV infection, and their loss has been shown to affect the gut microbial structure, for this study negatively, we found no association between CD4+ T cell count and viral load with the microbial composition. This is consistent with Russo et al. (Russo *et al*., 2022) found a microbiota with unchanged overall bacterial diversity in individuals with HIV-1 infection, before ART and after reaching virological suppression within 24 weeks of ART. In contrast, other studies reported an association of these factors with microbial diversity (Lu *et al*., 2018; Lyu *et al*., 2021). Large numbers of our study participants may be necessary to detect such signals. Further work would benefit from relating microbiome differences to more in-depth immune phenotyping of this population.

Our results are also consistent with another study conducted in Mexico that found that PI/r-based regimens had more severe effects than NNRTI-based regimens on gut microbiome diversity and composition (Villanueva-Millán *et al*., 2017). We identified lower alpha diversity and altered bacterial composition in individuals undergoing PI/r treatment compared to others on NNRTI regimens. Protease inhibitors are known to induce greater immunological restitution by an increased reduction in proviral DNA compared to NNRTI regimens (Llibre and Martínez-Picado, 2008; Villanueva-Millán *et al*., 2017; Ray *et al*., 2021).

### Study Limitations

Because the study HIV study participants received treatment with both ART and prophylactic cotrimoxazole, we cannot differentiate how these drugs impact diversity individually. Although it is tempting to speculate that these differences observed are driven by long-term antibiotic use, the multifactorial nature of HIV management makes it difficult to identify which disease or treatment aspects are driving the microbiome changes.

It is also possible that our limited sample size was underpowered to detect a difference where one existed. Given the complexity of the microbiome, we suggest future microbiota studies in HIV disease should be designed to minimise the influence of confounders and address environmental factors.

## Conclusion

In summary, our findings on the early impact on the gut microbiota in HIV-infected individuals initiating daily prophylaxis show a gradual reduction in diversity, with the most significant change after day 5. We also observed a composition shift characterised by the enrichment of species belonging to the *Prevetollea* and *Ruminococcus* genera and the depletion of species belonging to the Lactococcus and Bacteroides genera. These changes occur within five days and last well beyond 30 days post-initiation. This trend and significant shifts in functional microbial communities suggest an early onset of dysbiosis following TMP/SXT initiations. Further studies are needed to investigate the myriad of factors associated with HIV by leveraging a more robust multi-omics approach to offer a guide for future exploration of the effects of daily antibiotics during HIV disease.

## Data Availability

All data produced in the present study are available upon reasonable request to the authors

## Acknowledgements

We want to thank the staff of the Makerere Joint AIDS Project Clinic at Mulago Hospital: Moses M. Luutu and the team of the Molecular laboratory at Makerere University for technical support.

## Author contributions

CAA, CFN & BA conceived the study, and CAA and DPK designed the experiments. CAA performed experiments, and CAA, AM, KE & SM analysed the data. CAA, MA, and BA interpreted the data and wrote the paper. All authors reviewed and edited the manuscript.

## Availability of data and material

Raw sequences and study metadata generated during the study are available in the NCBI SRA repository https://www.ncbi.nlm.nih.gov/bioproject/.

